# Pulmonary function among rural residents in high air pollution area in northern Thailand

**DOI:** 10.1101/2022.05.16.22275174

**Authors:** Pitchayapa Ruchiwit, Narongkorn Saiphoklang, Kanyada Leelasittikul, Apiwat Pugongchai, Orapan Poachanukoon

**Affiliations:** Department of Medicine, Faculty of Medicine, Thammasat University, Pathum Thani, Thailand; Medical Diagnostics Unit, Thammasat University Hospital, Pathum Thani, Thailand; Center of Excellence for Allergy, Asthma and Pulmonary Diseases, Thammasat University Hospital, Pathum Thani, Thailand; Department of Pediatrics, Faculty of Medicine, Thammasat University, Pathum Thani, Thailand

**Author notes:** **Corresponding author:** (NS).

**Keywords:** air pollution, asthma, chronic obstructive pulmonary disease, northern, pulmonary functions, small airway disease, Thailand

## Abstract

**Background:** Air pollution has become a serious environmental and health issue in several countries. This condition leads to respiratory diseases, particularly asthma and chronic obstructive pulmonary disease (COPD). This study aimed to determine pulmonary functions and prevalence of respiratory diseases among rural residents in an area in northern Thailand with a high concentration of air pollution.

**Methods:** A cross-sectional study was conducted in people aged 18 years or older, living in Lamphun, Thailand in December 2021. Demographics, pre-existing diseases, respiratory symptoms, and pulmonary functions by spirometry including forced vital capacity (FVC), forced expiratory volume in one second (FEV_1_), peak expiration flow (PEF), forced expiration flow rate at 25-75% of FVC (FEF_25-75_), and bronchodilator responsiveness (BDR; FEV_1_ improvement after BDR test >12% and 200 mL) were collected.

**Results:** A total of 127 people (78.7%male) were included. Mean age was 43.76±11.32 years. Smoking was 52.0% and 4.44±5.45 pack-years. Self-reported respiratory diseases were allergic rhinitis (7.1%), asthma (0.8%), and COPD (0.8%). Respiratory symptoms were presented in 33.1% (14.2% runny nose, 10.2% nasal obstruction, 9.4% cough, 7.9% sputum production, and 6.3% breathlessness). Lung functions showed FVC in 96.74±12.91%, FEV_1_ in 97.52±12.99%, PEF in 102.46±19.18%, and FEF_25-75_ in 96.77±29.88%. Abnormal lung functions were found in 15.7%. Small airway disease (FEF_25-75_<65%) was 7.1%. Restrictive defect (FVC<80%) was 6.3%. Airway obstruction (FEV_1_/FVC<70%) was 2.4%. There was no BDR. Compared to people with normal lung functions, the abnormal lung function group was older (48.00±8.68 years vs 42.96±11.61 years, P=0.036), and had a higher proportion of breathlessness (20.0% vs 3.7%, P=0.021).

**Conclusions:** Abnormal pulmonary functions, especially small airway disease, were relatively common in rural residents in a polluted air area in northern Thailand. These abnormal pulmonary functions were associated with more respiratory symptoms.

**Clinicaltrials.in.th number:** TCTR20211223001

## Introduction

The problem of air pollution has become a serious environmental and health issue in many countries, including Thailand [1-3]. There are several small particles of air pollution, including sulfur dioxide (SO_2_), nitrogen dioxide (NO_2_), carbon monoxide (CO), ozone (O_3_), and particulate matter (PM), particularly fine particulate matter (PM_2.5_) and coarse particulate matter (PM_10_) [4, 5]. These are related to health problems such as increased mortality rate from lung cancer and cardiopulmonary disease [6, 7], and increased risk of respiratory diseases, especially chronic obstructive pulmonary disease (COPD) [4, 8], some interstitial lung diseases [9], asthma [10-13], and asthma exacerbation [14, 15]. Moreover, air pollution leads to decreased pulmonary functions [16-18], restricted activities [19], and increased respiratory symptoms, e.g., cough, sputum production, wheeze, breathlessness, and chest tightness [19].

Lamphun is a small province in northern Thailand that has been affected by air pollution because it is surround by mountains [20]. Especially in the summer season, air pollution is the main issue because low airflow causes accumulation of air pollutants. For the past 10 years, this problem in Lamphun has been monitored and found to be over the limit. Air pollution concentration exceeded the standard concentration level every dry season which affected human health [1]. However, pulmonary function data of people living in this area has been limited. Therefore, the objective of this study was to determine pulmonary functions and prevalence of respiratory diseases among rural residents in an area in northern Thailand with highly polluted air.

## Methods

### Study design and participants

A cross-sectional study was conducted at Li district, Lamphun in Thailand in December 2021. People aged 18 years or older and able to communicate in Thai were included in this study. Exclusion criteria were recent unstable angina or myocardial infarction, pulmonary embolus, resting heart rate more than 120 beats per minute, systolic blood pressure more than 180 mmHg or/and diastolic blood pressure more than 100 mmHg, inability to perform spirometry, and positive results of rapid antigen testing for SARS-CoV-2.

Ethic approval was obtained from the Human Research Ethics Committee of Thammasat University (Faculty of Medicine), Thailand (IRB No. MTU-EC-IM-6-295/64, COA No. 302/2021), in compliance with Declaration of Helsinki, The Belmont Report, CIOMS Guidelines and The International Practice (ICH-GCP). All methods were performed in accordance with these guidelines and regulations. All participants provided written informed consent.

### Procedures and outcomes

Demographic data, pre-existing diseases, respiratory symptoms, and pulmonary functions by spirometry including forced vital capacity (FVC), forced expiratory volume in one second (FEV_1_), peak expiration flow (PEF), forced expiration flow rate at 25-75% of FVC (FEF_25-75_), and bronchodilator responsiveness (BDR) were collected. Spirometry was performed according to the American Thoracic Society and European Respiratory Society guidelines [21-23] using PC spirometer (Vyntus SPIRO, Vyaire Medical, Inc., Mettawa, IL, USA). Briefly, participants were asked to blow into the tube hard and fast and then to continue exhaling for 15 seconds or more. FVC, FEV_1_, FEV_1_/FVC, PEF and FEF_25-75_ were recorded and reported in liters (L), %predicted, %, or liters per second (L/s). BDR testing was done using salbutamol inhalation (total dose of 400 µg) and repeating spirometry 15 minutes later according to the American Thoracic Society and European Respiratory Society guidelines [21-23].

Abnormal lung functions were defined as airway obstruction (FEV_1_/FVC<70%), restrictive lung disease (FVC<80%), small airway disease (FEF_25-75_<65%), or BDR (FEV_1_ improvement after BDR test >12% and 200 mL).

### Statistical analysis

In a previous study [24], the prevalence of airway obstruction in subjects living in an outdoor air pollution area in India was 24.9%. Our sample size was calculated using 80% power, 5% type I error, and 8% precision margin. Thus, the sample size would be 113.

Categorical data was shown as number (%). Continuous data was shown as mean ± standard deviation. Chi-squared test was used to compare categorical variables between two groups. Student’s t-test was used to compare continuous variables between two groups. A two-sided p-value <0.05 was considered statistically significant. Statistical analyses were performed using SPSS version 26.0 software.

## Results

A total of 140 subjects were screened. Of these, 127 were included in the study and 23 were excluded (Figure 1). 78.7% was male. Mean age was 43.76±11.32 years. Current or former smokers comprised 52.0% and 4.44±5.45 pack-years. The most common occupation was government officer (45.7%). Biomass fuel was used for 51.2% of cooking. Allergic rhinitis (7.1%) was the most common self-reported respiratory disease. Runny nose (14.2%) was the most common presenting respiratory symptom (Table 1).

**Fig 1.**
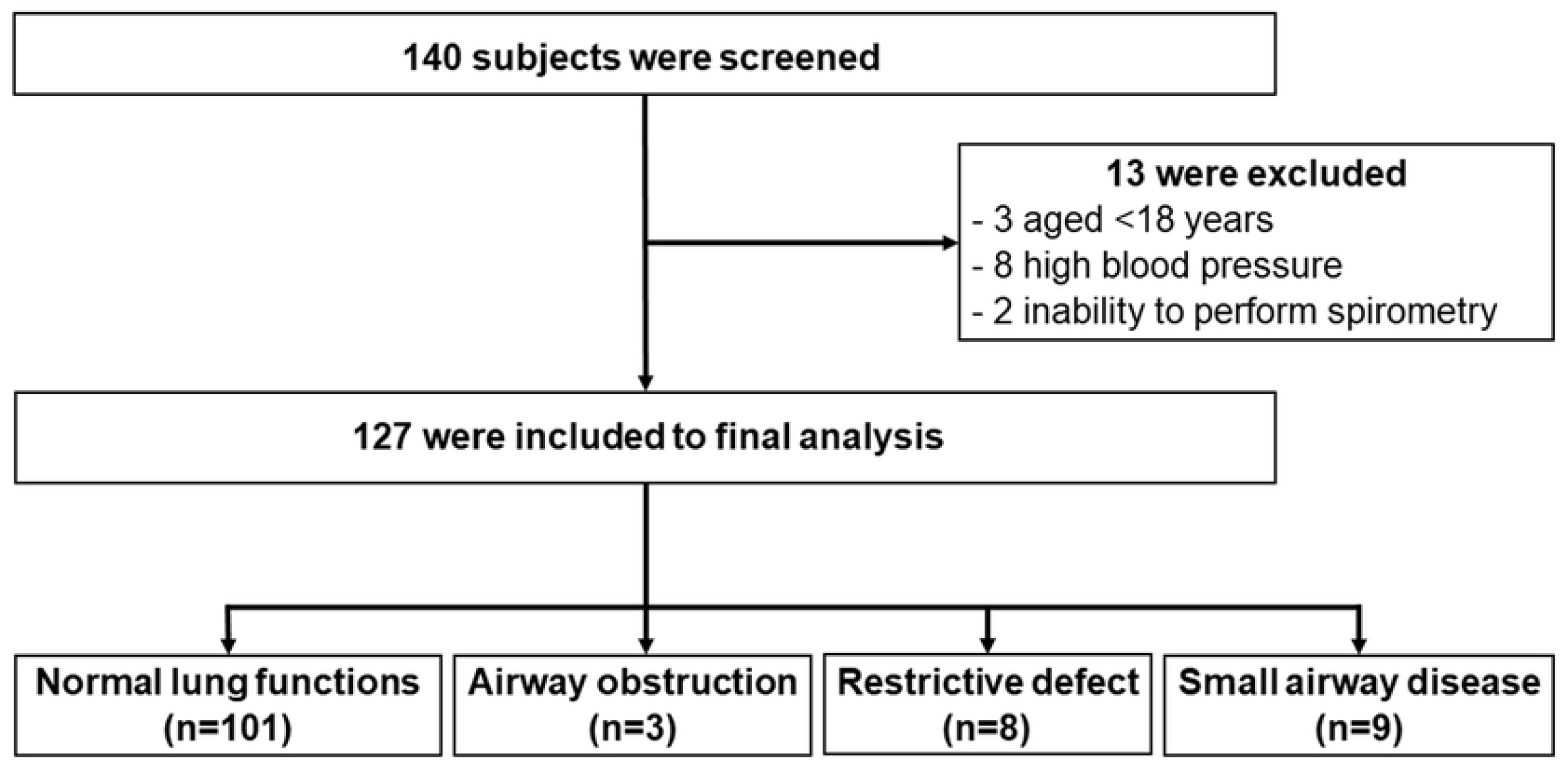
Flowchart of subject recruitment to the study.

**Table 1.** Baseline characteristics of subjects living in air pollution area.

| Characteristics | Data (n=127) |
| --- | --- |
| Age, years | $43.76 \pm 11.32$ |
| Male | 100 (78.7) |
| Body mass index, kg/m<sup>2</sup> | 24.55±4.24 |
| Smoking (current or former) | 66 (52.0) |
| Amount of smoking, pack-years | 4.44±5.45 |
| Working, hours/week | 39.31±9.85 |
| **Occupations** |  |
| Government officer | 58 (45.7) |
| Farmer | 41 (32.3) |
| General employee | 16 (12.6) |
| Others | 9 (7.0) |
| Unemployed | 3 (2.4) |
| **Comorbidities** |  |
| Hypertension | 24 (18.9) |
| Dyslipidemia | 14 (11.0) |
| Diabetes | 10 (7.9) |
| Obesity | 1 (0.8) |
| **Risks of respiratory diseases and protection** |  |
| Outdoor job | 86 (67.7) |
| Biomass fuel used for cooking | 65 (51.2) |
| Mask worn for smog protection | 90 (70.9) |
| **Self-reported respiratory diseases** |  |
| Allergic rhinitis | 9 (7.1) |
| Asthma | 1 (0.8) |
| Chronic obstructive pulmonary disease | 1 (0.8) |
| **Respiratory symptoms** | 42 (33.1) |
| Runny nose | 18 (14.2) |
| Nasal obstruction | 13 (10.2) |
| Cough | 12 (9.4) |
| Sputum production | 10 (7.9) |
| Breathlessness | 8 (6.3) |
| Chest tightness | 4 (3.1) |
| Sore throat | 4 (3.1) |
| Wheezes | 2 (1.6) |

Data shown as n (%) or mean±SD

Abnormal lung functions were found in 15.7% of subjects. Airway obstruction was 2.4%. Restrictive defect was 6.3%. Small airway disease was 7.1%. There was no BDR (Table 2). Compared to people with normal lung functions, the abnormal lung function group was significantly older, and had a higher proportion of breathlessness (Table 3). The dataset of study participants is shown in S1 File.

**Table 2.** Lung function data of subjects living in air pollution area.

| Parameters | Data (n=127) |
| --- | --- |
| FVC, L | 3.56±0.75 |
| FVC, %predicted | 96.74±12.9 |
| FEV<sub>1</sub>, L | 2.97±0.65 |
| FEV<sub>1</sub>, % predicted | 97.52±12.99 |
| FEV<sub>1</sub> improvement after BDR test, % | 1.92±4.13 |
| FEV<sub>1</sub>/FVC, % | 83.67±5.81 |
| PEF, L/s | 8.10±1.85 |
| PEF, % predicted | 102.46±19.18 |
| FEF<sub>25-75</sub>, L/s | 3.28±1.17 |
| FEF<sub>25-75</sub>, %predicted | 96.77±29.88 |
| Abnormal lung function | 20 (15.7) |
| FEV<sub>1</sub>/FVC <70% | 3 (2.4) |
| FVC <80% | 8 (6.3) |
| FEF<sub>25-75</sub><65% | 9 (7.1) |
| FEV<sub>1</sub> improvement after BD test >12% and 200 mL | 0 (0) |

Data shown as n (%) or mean±SD
BD=bronchodilator responsiveness, FEV<sub>1</sub>=forced expiratory volume in 1 second,
FVC=forced vital capacity, FEF<sub>25-75</sub>=forced expiratory flow at 25-75% of FVC, PEF=peak expiratory flow, L=liters, mL=milliliters, s=second

**Table 3.** Comparison in baseline characteristics of subjects living in air pollution area between the abnormal and the normal lung function groups.

| **Variables** | **Abnormal lung**<br>**function**<br>**(n=20)** | **Normal lung**<br>**function**<br>**(n=107)** | **P-value** |
| --- | --- | --- | --- |
| Age, years | 48.00±8.68 | 42.96±11.61 | 0.036 |
| Male | 15 (75.0) | 85 (79.4) | 0.766 |
| Body mass index, kg/m<sup>2</sup> | 25.00±4.44 | 24.47±4.22 | 0.628 |
| Smoking | 10 (50.0) | 56 (52.3) | 0.848 |
| Smoking, pack-years | 2.78±1.91 | 4.80±5.91 | 0.381 |
| Working, hours/week | 37.50±6.83 | 39.61±10.28 | 0.429 |
| Outdoor job | 12 (60.0) | 74 (69.2) | 0.231 |
| Mask worn for smog protection | 17 (89.5) | 73 (70.2) | 0.081 |
| Biomass fuel used for cooking | 8 (40.0) | 57 (56.4) | 0.178 |
| Hypertension | 6 (30.0) | 18 (16.8) | 0.211 |
| Dyslipidemia | 5 (25.0) | 9 (8.4) | 0.046 |
| Diabetes | 1 (5.0) | 9 (8.4) | 1.000 |
| Obesity | 0 (0) | 1 (0.9) | 1.000 |
| Allergic rhinitis | 1 (5.0) | 8 (7.5) | 1.000 |
| Asthma | 0 (0) | 1 (0.9) | 1.000 |
| Chronic obstructive pulmonary disease | 1 (5.0) | 0 (0) | 0.157 |
| Presence of respiratory symptom | 8 (40.0) | 34 (31.8) | 0.473 |
| Runny nose | 4 (20.0) | 14 (13.1) | 0.483 |
| Nasal obstruction | 2 (10.0) | 11 (10.3) | 1.000 |
| Cough | 1 (5.0) | 11 (10.3) | 0.690 |
| Sputum production | 0 (0) | 10 (9.3) | 0.361 |
| Breathlessness | 4 (20.0) | 4 (3.7) | 0.021 |
| Chest tightness | 0 (0) | 4 (3.7) | 1.000 |
| Sore throat | 0 (0) | 4 (3.7) | 1.000 |
| Wheezes | 1 (5.0) | 1 (0.9) | 0.291 |

Data shown as n (%) or mean±SD

## Discussion

This study is the first study of pulmonary function and prevalence of respiratory diseases in a high air pollution area in northern Thailand. Abnormal lung function was found in 15.7% of subjects. The most common abnormality was small airway disease (7.1%). Airway obstruction was found in only 2.4% of the subjects in our study, which is 10-fold less than the prevalence in people living in an outdoor air pollution area in northern India (24.9%), according to a study by Kumar R and colleagues [24]. Those findings might be because air pollution levels, particularly PM_2.5_, are significantly higher in in northern India than in northern Thailand. High PM_2.5_ concentration in northern India is a major problem, which is associated with social disadvantages [25]. The main source of PM_2.5_ in India is the post-harvest agricultural fire activity [26].

The north of Thailand is faced with high air pollution because the natural geographical features of this area are mountains and basin pan areas [27]. Forest fires and the burning of agricultural waste are common causes of air pollution in this area [2, 28, 29]. Additionally, pollution might blow in from neighboring countries as transboundary pollution [29]. These sources all contribute to the accumulation of pollutants [1, 29, 30]. It usually from late winter to early spring because the weather is calm and dry [1, 4, 29]. Small air pollutants cannot float to the atmosphere, so they accumulate in this condition.

The Pollution Control Department, Thailand, reported that in 2021, the year of our study, the mean PM_2.5_ level across the country was 35.11 μg/m^3^ (range in 2 to128 μg/m^3^) while the mean PM_10_ level was 47.28 μg/m3 (range in 10 to152 μg/m^3^) [31]. These levels are higher than the World Health Organization’s recommended annual average PM_2.5_ and PM_10_ concentrations of 5.0 and 15 μg/m^3^, respectively [5]. The results of our study indicate that air pollution affects the health of residents in northern Thailand.

Numerous pollutants, especially PM_2.5_ and PM_10_, are associated with health problems. Several previous studies found that PM_2.5_ led to decreased pulmonary functions (FVC, FEV_1_ and maximal mid-expiratory flow) [32, 33], increased risk of respiratory diseases and symptoms [34], airway inflammation, and lung fibrosis [35-37]. Decreased maximal mid-expiratory flow, or FEF_25-75_<65%, a small airway disease, was a common abnormal pulmonary function in our study (7.1%).

A large study from results of the population-based UK Biobank study by Doiron D and colleagues showed that ambient air pollution was associated with lower lung functions (FVC and FEV_1_) and increased COPD prevalence especially high concentrations of PM_2.5_, PM_10_ and NO_2_ [38]. Furthermore, the study of air pollution from forest fire in northern California, the United States, by Reid CE and workers revealed that every 5 μg/m^3^ increase in PM_2.5_ increased the risk of asthma exacerbation requiring hospitalization (relative risk of 1.07) or emergency department (ED) visits (relative risk of 1.06), or COPD exacerbation with ED visits (relative risk of 1.02) [39].

A systematic review of acute outdoor air pollution on PEF in patients with asthma by Edginton S and coworkers showed that each 10 μg/m^3^ increase in acute exposure to PM_10_ and PM_2.5_ was associated with a decrease in PEF of 0.25 L/min in adults with asthma, particularly among non-smokers [40]. In addition, every 10 μg/m^3^ increase in PM_2.5_ increased the risk of COPD exacerbation requiring ED visits or hospitalizations by 2.5% [41].

A systematic review of short-term exposure to air pollution by Achilleos S and colleagues showed that every 10 μg/m^3^ increase in PM_2.5_ increased mortality rates from all causes by 0.89%, cardiovascular diseases by 0.80%, and respiratory diseases by 1.10% [42]. Moreover, several studies of long-term exposure to ambient air pollution showed every 10 μg/m^3^ increase in PM_2.5_ increased mortality rates from all causes by 4-6% [43, 44], cardiac diseases by 11-14% [44, 45], and lung cancer by 8-21% [43, 45], and it decreased pulmonary function (decline of FEV_1_ in 3.38 mL per increase 10 μg/m^3^ of PM_10_) [46].

Our study found that the prevalence of abnormal lung function was not very high, probably because the mean age of participants was only 43 years and the associated risk factor smoking only 4.4 pack-years. These findings are similar to a study of lung functions among university athletes by Saiphoklang N and coworkers, which showed that young athletes who smoked very lightly had good lung functions [47].

Our study showed that subjects with abnormal lung functions were significantly older than those with normal lung functions. This finding indicates that older age and long-term exposure to air pollutants might be risk factors for lung diseases, particularly COPD, which is similar to findings of some other studies [48, 49]. Moreover, our study also showed that abnormal lung functions were associated with breathlessness.

Interestingly, a study of children (median age of 7.7 years) with long-term exposure to air pollution by Rice MB and colleagues showed that this situation, particularly living less than 100 m from a major roadway, was associated with lower lung function in terms of FVC particularly living less than 100 m from a major roadway [50]. However, the mechanism of pollution-induced lung damage is unclear. Some studies suggested that PM_2.5_ and PM_10_ might produce adverse effects on the respiratory system via the generation of reactive oxygen species [35, 36] lead to airway inflammation and changes in pulmonary function [36].

There are the limitations of this study. Firstly, because of the small population size, some results might not be obviously different between the abnormal and normal lung function groups. Secondly, our study is a cross-sectional study. Subjects were not followed up in long-term to monitor lung functions and respiratory symptoms. Therefore, we cannot predict the changes of symptoms and lung function in the future. However, our study showed a negative trend of obstructive defect. Large prospective studies are required to evaluate lung functions and long-term clinical outcomes in respiratory diseases in northern Thailand.

## Conclusions

Abnormal pulmonary functions, especially small airway disease, were relatively common in rural residents in an air pollution area in northern Thailand. These abnormal pulmonary functions were associated with more respiratory symptoms, particularly breathlessness. Large prospective studies are needed to investigate lung functions and long-term clinical outcomes in respiratory diseases in this area.

## Data Availability

All relevant data are within the manuscript and its Supporting Information files.

## Supporting information

**S1 File. Dataset of study participants**.

## Acknowledgments

The authors would like to thank Michael Jan Everts, Faculty of Medicine, Thammasat University, for proofreading this manuscript.

This work was supported by the Center of Excellence for Allergy Asthma and Pulmonary Diseases (TU-CAAP), the Medical Diagnostics Unit (MDU), Thammasat University Hospital, and the Research Group in Airway Diseases and Allergy, Faculty of Medicine, Thammasat University, Thailand.

## References

1. Kliengchuay W, Worakhunpiset S, Limpanont Y, Meeyai AC, Tantrakarnapa K. Influence of the meteorological conditions and some pollutants on PM10 concentrations in Lamphun, Thailand. J Environ Health Sci Eng. 2021;19:237–49.

2. Phairuang W, Suwattiga P, Chetiyanukornkul T, Hongtieab S, Limpaseni W, Ikemori F, et al. The influence of the open burning of agricultural biomass and forest fires in Thailand on the carbonaceous components in size-fractionated particles. Environ Pollut. 2019;247:238–47.

3. Kliengchuay W, Cooper Meeyai A, Worakhunpiset S, Tantrakarnapa K. Relationships between Meteorological Parameters and Particulate Matter in Mae Hong Son Province, Thailand. Int J Environ Res Public Health. 2018;15.

4. Wang F, Ni SS, Liu H. Pollutional haze and COPD: etiology, epidemiology, pathogenesis, pathology, biological markers and therapy. J Thorac Dis. 2016;8:E20–30.

5. World Health Organization. Ambient (outdoor) air pollution 2021 [updated April 22, 2022. September 22, 2021:[Available from: https://www.who.int/news-room/fact-sheets/detail/ambient-(outdoor)-air-quality-and-health.

6. Dockery DW, Pope CA, 3rd, Xu X, Spengler JD, Ware JH, Fay ME, et al. An association between air pollution and mortality in six U.S. cities. N Engl J Med. 1993;329:1753–9.

7. Pope CA, 3rd, Burnett RT, Thurston GD, Thun MJ, Calle EE, Krewski D, et al. Cardiovascular mortality and long-term exposure to particulate air pollution: epidemiological evidence of general pathophysiological pathways of disease. Circulation. 2004;109:71–7.

8. Shin S, Bai L, Burnett RT, Kwong JC, Hystad P, van Donkelaar A, et al. Air pollution as a risk factor for incident chronic obstructive pulmonary disease and asthma. A 15-year population-based cohort study. Am J Respir Crit Care Med. 2021;203:1138–48.

9. Singh N, Singh S. Interstitial lung diseases and air pollution: Narrative review of literature. Pulm Ther. 2021;7:89–100.

10. Sio YY, Chew FT. Risk factors of asthma in the Asian population: a systematic review and meta-analysis. J Physiol Anthropol. 2021;40:22.

11. Liu S, Jorgensen JT, Ljungman P, Pershagen G, Bellander T, Leander K, et al. Long-term exposure to low-level air pollution and incidence of asthma: the ELAPSE project. Eur Respir J. 2021;57.

12. Liu S, Lim YH, Pedersen M, Jorgensen JT, Amini H, Cole-Hunter T, et al. Long-term exposure to ambient air pollution and road traffic noise and asthma incidence in adults: The Danish Nurse cohort. Environ Int. 2021;152:106464.

13. Zhang Y, Wei J, Shi Y, Quan C, Ho HC, Song Y, et al. Early-life exposure to submicron particulate air pollution in relation to asthma development in Chinese preschool children. J Allergy Clin Immunol. 2021;148:771–82 e12.

14. Huang J, Yang X, Fan F, Hu Y, Wang X, Zhu S, et al. Outdoor air pollution and the risk of asthma exacerbations in single lag0 and lag1 exposure patterns: a systematic review and meta-analysis. J Asthma. 2021:1–18.

15. Khatri SB, Newman C, Hammel JP, Dey T, Van Laere JJ, Ross KA, et al. Associations of air pollution and pediatric asthma in Cleveland, Ohio. ScientificWorldJournal. 2021;2021:8881390.

16. He QQ, Wong TW, D. L, Jiang ZQ, Gao Y, Qiu H, et al. Effects of ambient air pollution on lung function growth in Chinese schoolchildren. Respir Med. 2010;104:1512–20.

17. Roy A, Hu W, Wei F, Korn L, Chapman RS, Zhang JJ. Ambient particulate matter and lung function growth in Chinese children. Epidemiology. 2012;23:464–72.

18. Frampton MW, Balmes JR, Bromberg PA, Arjomandi M, Hazucha MJ, Thurston SW, et al. Effects of short-term increases in personal and ambient pollutant concentrations on pulmonary and cardiovascular function: A panel study analysis of the Multicenter Ozone Study in oldEr subjects (MOSES 2). Environ Res. 2022;205:112522.

19. Karakatsani A, Analitis A, Perifanou D, Ayres JG, Harrison RM, Kotronarou A, et al. Particulate matter air pollution and respiratory symptoms in individuals having either asthma or chronic obstructive pulmonary disease: a European multicentre panel study. Environ Health. 2012;11:75.

20. Lamphun [Internet]. Wikipedia. 2022 [cited April 22, 2022]. Available from: https://en.wikipedia.org/wiki/Lamphun.

21. Miller MR, Crapo R, Hankinson J, Brusasco V, Burgos F, Casaburi R, et al. General considerations for lung function testing. Eur Respir J. 2005;26:153–61.

22. Miller MR, Hankinson J, Brusasco V, Burgos F, Casaburi R, Coates A, et al. Standardisation of spirometry. Eur Respir J. 2005;26:319–38.

23. Graham BL, Steenbruggen I, Miller MR, Barjaktarevic IZ, Cooper BG, Hall GL, et al. Standardization of spirometry 2019 update. An official American Thoracic Society and European Respiratory Society technical statement. Am J Respir Crit Care Med. 2019;200:e70–e88.

24. Kumar R, Sharma M, Srivastva A, Thakur JS, Jindal SK, Parwana HK. Association of outdoor air pollution with chronic respiratory morbidity in an industrial town in northern India. Arch Environ Health. 2004;59:471–7.

25. Iwanowicz-Palus G, Zarajczyk M, Bien A, Korzynska-Pietas M, Krysa J, Rahnama-Hezavah M, et al. The Relationship between Social Support, Self-Efficacy and Characteristics of Women with Diabetes during Pregnancy. Int J Environ Res Public Health. 2021;19.

26. Jethva H, Torres O, Field RD, Lyapustin A, Gautam R, Kayetha V. Connecting crop productivity, residue fires, and air quality over northern India. Sci Rep. 2019;9:16594.

27. Wikipedia. Geography of Thailand 2022 [cited 2022 April 20]. Available from: https://en.wikipedia.org/wiki/Geography_of_Thailand.

28. Vichit-Vadakan N, Vajanapoom N. Health impact from air pollution in Thailand: current and future challenges. Environmental health perspectives. 2011;119:A197–A8.

29. Chansuebsri S, Kraisitnitikul P, Wiriya W, Chantara S. Fresh and aged PM2.5 and their ion composition in rural and urban atmospheres of Northern Thailand in relation to source identification. Chemosphere. 2022;286:131803.

30. Vichit-Vadakan N, Vajanapoom N. Health impact from air pollution in Thailand: current and future challenges. Environ Health Perspect. 2011;119:A197–8.

31. Pollution Control Department. Air4Thai 2022 [cited 2022 April 25]. Available from: http://air4thai.pcd.go.th/webV3/#/Home.

32. Peters JM, Avol E, Gauderman WJ, Linn WS, Navidi W, London SJ, et al. A study of twelve Southern California communities with differing levels and types of air pollution. II. Effects on pulmonary function. Am J Respir Crit Care Med. 1999;159:768–75.

33. Guo C, Hoek G, Chang LY, Bo Y, Lin C, Huang B, et al. Long-term exposure to ambient fine particulate matter (PM2.5) and lung function in children, adolescents, and young adults: A longitudinal cohort study. Environ Health Perspect. 2019;127:127008.

34. Wei Y, Wang Y, Di Q, Choirat C, Wang Y, Koutrakis P, et al. Short term exposure to fine particulate matter and hospital admission risks and costs in the Medicare population: time stratified, case crossover study. BMJ. 2019;367:l6258.

35. Hogervorst JG, de Kok TM, Briede JJ, Wesseling G, Kleinjans JC, van Schayck CP. Relationship between radical generation by urban ambient particulate matter and pulmonary function of school children. J Toxicol Environ Health A. 2006;69:245–62.

36. Janssen NA, Strak M, Yang A, Hellack B, Kelly FJ, Kuhlbusch TA, et al. Associations between three specific a-cellular measures of the oxidative potential of particulate matter and markers of acute airway and nasal inflammation in healthy volunteers. Occup Environ Med. 2015;72:49–56.

37. Sun B, Shi Y, Li Y, Jiang J, Liang S, Duan J, et al. Short-term PM2.5 exposure induces sustained pulmonary fibrosis development during post-exposure period in rats. J Hazard Mater. 2020;385:121566.

38. Doiron D, de Hoogh K, Probst-Hensch N, Fortier I, Cai Y, De Matteis S, et al. Air pollution, lung function and COPD: results from the population-based UK Biobank study. Eur Respir J. 2019;54.

39. Reid CE, Jerrett M, Tager IB, Petersen ML, Mann JK, Balmes JR. Differential respiratory health effects from the 2008 northern California wildfires: A spatiotemporal approach. Environ Res. 2016;150:227–35.

40. Edginton S, O’Sullivan DE, King WD, Lougheed MD. The effect of acute outdoor air pollution on peak expiratory flow in individuals with asthma: A systematic review and meta-analysis. Environ Res. 2021;192:110296.

41. DeVries R, Kriebel D, Sama S. Outdoor air pollution and COPD-related emergency department visits, hospital admissions, and mortality: A meta-analysis. COPD. 2017;14:113–21.

42. Achilleos S, Kioumourtzoglou MA, Wu CD, Schwartz JD, Koutrakis P, Papatheodorou SI. Acute effects of fine particulate matter constituents on mortality: A systematic review and meta-regression analysis. Environ Int. 2017;109:89–100.

43. Pope CA, 3rd, Burnett RT, Thun MJ, Calle EE, Krewski D, Ito K, et al. Lung cancer, cardiopulmonary mortality, and long-term exposure to fine particulate air pollution. JAMA. 2002;287:1132–41.

44. Hoek G, Krishnan RM, Beelen R, Peters A, Ostro B, Brunekreef B, et al. Long-term air pollution exposure and cardio-respiratory mortality: a review. Environ Health. 2013;12:43.

45. Chen H, Goldberg MS, Villeneuve PJ. A systematic review of the relation between long-term exposure to ambient air pollution and chronic diseases. Rev Environ Health. 2008;23:243–97.

46. Heinrich J, Schikowski T. COPD patients as vulnerable subpopulation for exposure to ambient air pollution. Curr Environ Health Rep. 2018;5:70–6.

47. Saiphoklang N, Poachanukoon O, Soorapan S. Smoking characteristics and lung functions among university athletes. Sci Rep. 2020;10:20118.

48. Ackermann-Liebrich U, Leuenberger P, Schwartz J, Schindler C, Monn C, Bolognini G, et al. Lung function and long term exposure to air pollutants in Switzerland. Study on Air Pollution and Lung Diseases in Adults (SAPALDIA) Team. Am J Respir Crit Care Med. 1997;155:122–9.

49. Elbarbary M, Oganesyan A, Honda T, Kelly P, Zhang Y, Guo Y, et al. Ambient air pollution, lung function and COPD: cross-sectional analysis from the WHO Study of AGEing and adult health wave 1. BMJ Open Respir Res. 2020;7.

50. Rice MB, Rifas-Shiman SL, Litonjua AA, Oken E, Gillman MW, Kloog I, et al. Lifetime exposure to ambient pollution and lung function in children. Am J Respir Crit Care Med. 2016;193:881–8.

